# Integration of tissue-specific multi-omics data implicates brain targets for complex neuropsychiatric traits

**DOI:** 10.1101/2023.06.14.23291366

**Authors:** Shan Cong, Zhiling Sang, Luolong Cao, Junbo Yuan, Yanhong Li, Hong Liang, Xiaohui Yao

**Author notes:** **Correspondence to:** Xiaohui Yao, 423 11# Building, 1777 Sansha Road, Qingdao, Shandong 266000, China.

## Abstract

Genome-wide association studies (GWAS) have uncovered genetic variants susceptible to brain disorders. However, due to the complex pathogenesis of these diseases and heterogeneity of the brain tissues, how and through which the genetic variants confer risk for brain abnormalities and brain disorders remain elusive, especially from a multi-omics perspective and in the context of brain regions. In this study, we integrated brain region-specific transcriptomics, proteomics, and imaging genetics data by systematically applying transcriptome- and proteome-wide association analysis, Mendelian randomization, and Bayesian colocalization methods. At both gene expression and protein abundance levels, this integrative study identified 51 associations linking 42 targets to structural alterations of 10 brain regions. Additionally, we validated the causal effects of 20 identified genes on one or more brain disorders. Our analysis further illuminated the significant enrichment of 12 targets in five main types of brain cells. Overall, this study underscored the utility of a multi-omics and region-specific approach in understanding the pathogenesis of complex brain abnormalities and brain disorders.

## INTRODUCTION

Brain imaging biomarkers significantly contribute to early diagnosis of numerous brain disorders, such as Alzheimer’s disease (AD) where brain changes can be observed years before clinical symptoms arise [1]. Advancements in high-throughput sequencing and imaging technologies have fostered the emergence of brain imaging genetics, a field exploring the impact of genetic variants on brain imaging quantitative traits (iQTs). Genome-wide association studies (GWAS) of brain iQTs have identified numerous susceptible genetic variants [2]. However, the molecular mechanisms underlying brain tissues and brain diseases are very complicated and cannot be exhaustively explained only from the genetics perspective. Many genetic variants exert effects by regulating gene expression and protein abundance, moreover, often in a tissue-specific manner. Yet, the cost and invasive nature of obtaining brain-specific omics data (e.g., transcriptomics and proteomics) presents challenges, especially for conducting large-scale studies in the context of brain disorders.

To address the aforementioned problem, recent efforts have been made to develop computational and statistical models to estimate the effects of gene expression on disease, by integrating expression quantitative trait locus (eQTL) data derived from small-size reference panel and summary statistics from large-scale GWAS. Transcriptome-wide association study (TWAS) [3] and Mendelian randomization (MR) [4] are two main types of approaches utilized extensively to discover causal genes implicated in complex diseases. A few studies have applied TWAS and/or MR methods as well as their variants and have reported numerous causal genes for complex diseases and traits, particularly brain disorders. For example, several studies [5–7] applied individual- or summary-based TWAS/MR approaches and identified a number of AD risk genes with brain regional specificity. Restuadi et al. [8] evaluated the effect of gene expression on Amyotrophic Lateral Sclerosis (ALS) using both summary-based MR (SMR) and TWAS, and identified *GPX3* and *TNIP1*. Yang et al. [9] used the SMR approach and identified multiple genes across several brain regions with pleiotropic associations with major depressive disorder (MDD). Recently, research efforts [10,11] have expanded from mRNA expression to protein abundance to discover therapeutic targets for brain diseases at the proteomic level.

Few integrative studies have focused on investigating causal molecules for brain iQTs, and prior studies are mainly dedicated on exploring the molecular relevance within gene expression level. For example, Zhao et al. [12] employed UTMOST [13] to perform a cross-tissue TWAS analysis of 211 brain structural iQTs and reported 918 significant gene-iQT associations. Similarly, Mai et al. [14] conducted S-PrediXcan [15], a TWAS method, to integrate brain tissue-specific transcriptomics data with brain imaging GWAS summary statistics for total brain volume and intracranial volume and discovered 10 associated genes. However, to our knowledge, no study has examined proteins associated with brain iQTs, despite mRNA and protein complementing each other and being both necessary for comprehensive brain understanding.

To fill this gap, we aimed to uncover brain biomarkers and reveal the molecular mechanisms underpinning brain from a multi-omics perspective and in the context of brain tissues. We integrated brain tissue-specific transcriptomics, proteomics, and imaging genetics data by systematically applying TWAS, PWAS, MR and Bayesian colocalization. Particularly, we integrated brain structural meta-GWAS results from the Enhancing Neuroimaging Genetics through Meta Analysis (ENIGMA) consortium [16] with brain-derived QTLs and molecular weights computed from five cohorts, including the Religious Orders Study and Memory and Aging Project (ROSMAP), Mayo RNAseq Study (Mayo), NIMH Human Brain Collection Core (HBCC), the CommonMind Consortium (CMC) and MSBB for discovery. Genetic colocalization pointed towards the significant shared genetic underpinning between gene transcription and protein synthesis in the brain. Brain single-cell expression data was employed to examine the enrichment of identified genes in brain cell types. Finally, we validated the causal effects of candidates on brain disorders using GWAS summary statistics for five brain diseases. Figure 1 provides an overview of the analytical pipeline employed in our study.

**Figure 1.**
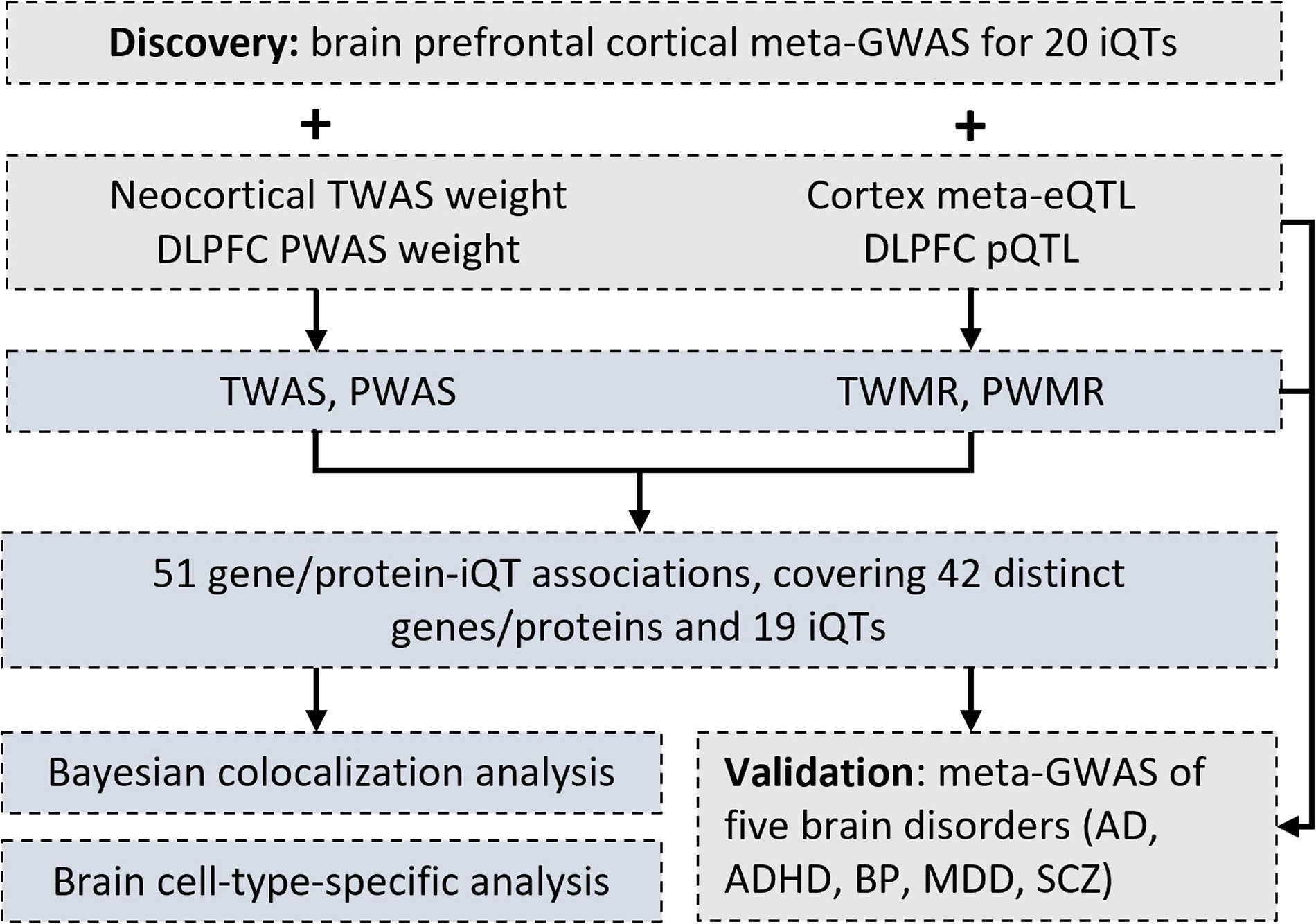
Flowchart illustrating the design of the brain tissue-specific multiomics integration study.

## MATERIALS AND METHODS

### Brian-derived QTLs and weights data

We used four brain-derived datasets delineating relationships between genetic variants and corresponding genes/proteins, including meta-eQTL of the brain cortex, gene expression weights calculated from the neocortical region, pQTLs estimated from the dorsolateral prefrontal cortex (DLPFC), and protein abundance weights calculated from the DLPFC. Subsequent sections provide a detailed description of each dataset.

#### Cortex meta-eQTL

The eQTL data were sourced from a large-scale brain eQTL meta-analysis (*N*=1 433) [17]. Briefly, Sieberts et al. first generated cerebral cortical eQTL data from four individual cohorts, including ROSMAP and Mayo RNAseq Study (Mayo) from the Accelerating Medicines Partnership for Alzheimer’s Disease (AMP-AD) Consortium, and MSSM-Penn-Pitt study and NIMH Human Brain Collection Core (HBCC) from the CommonMind Consortium (CMC). They subsequently performed a meta-analysis using cortical eQTL results derived from ROSMAP DLPFC, Mayo temporal cortex, MSSM-Penn-Pitt and HBCC. We downloaded the meta-eQTL summary statistics from Synapse (https://doi.org/10.7303/syn16984815.1). This comprehensive *cis*-eQTL data included 100 080 618 SNP-gene expression pairs. A significant threshold of *P*-value < 5E-8 was employed, reducing it to 1 403 800 *cis*-eQTLs included in the MR analysis.

#### Neocortical gene expression weight

The TWAS weights across multiple neocortical regions were derived from the combined ROSMAP, Mayo, and MSBB cohorts [5]. A training dataset was constructed comprising 790 individual genotype profiles paired with 888 RNA-Sequencing (RNA-Seq) samples originated from 6 neocortical regions: temporal cortex, prefrontal cortex, inferior frontal gyrus, superior temporal gyrus, parahippocampal cortex, and DLPFC. The FUSION software [3] was tailored to accommodate multiple RNA-Seq profiles from different regions of a single individual, by grouping all samples from a given individual into a single cross-validation fold during the training and optimization process. Five TWAS models (top1, blup, lasso, enet and bslmm) were utilized to train and yield SNP weights for each gene. We retrieved the TWAS weights from Synapse (https://doi.org/10.7303/syn2580853) that included a total of 6 780 trained weights in the form of RData.

#### DLPFC pQTL

Wingo et al. [18] employed a linear regression model, as implemented in Plink [19], to estimate *cis*-pQTLs from the DLPFC proteomes and corresponding genotyping data for 376 individuals in the ROSMAP study. To reduce the effects of linkage disequilibrium (LD), only SNPs present in an LD reference panel provided by FUSION were considered in the analysis. After preprocessing and quality control, 8 356 proteins and 1 190 321 SNPs were retained. Detailed information about the preprocessing and quality control steps for both the proteomics and genetics data is available in [18]. The summary statistics of the ROSMAP pQTL can be accessed at Synapse (https://doi.org/10.7303/syn23627957), including a total of 928 865 SNP-protein pairs. For our analysis, we applied a significant level of *P* < 5E-8, resulting a total of 14 221 pQTLs incorporated into the MR analysis.

#### DLPFC protein expression weight

Protein expression weights specific to the DLPFC were also derived from the ROSMAP study [18]. As mentioned above, 376 subjects with matched DLPFC proteome profiles and genotyping data were included. The heritability for each protein was quantified using both proteomics and genetics data. Proteins exhibiting significant heritability (*P* < 0.01) were investigated further, by applying the FUSION approach to estimate the influence of genetic variants on protein abundance. Five predictive models (top1, blup, lasso, enet and bslmm) were employed, and protein weights from the model that yielded the highest predictive value were selected. The trained weights were obtained from Synapse (https://doi.org/10.7303/syn23627957), with a total of 1 761 weight files in RData format. For more detailed information, please refer to [18].

### GWAS summary statistics data

We used GWAS summary statistics of brain imaging traits to infer risk molecular markers associated with brain tissues. The findings were subsequently validated by evaluating their causal effects on five brain disorders.

#### GWAS of brain structural iQTs

We employed summary meta-GWAS statistics of human cortical structures from the ENIGMA-3 study conducted by Grasby et al. [16]. This study executed a principal meta-GWAS on brain magnetic resonance imaging (MRI) data from 33 992 participants of European ancestry. In brief, the surface area (SA) and averaged thickness (aTH) of 34 individual cortical regions, as defined by the Desikan-Killiany (DK) atlas, were extracted from the MRI scans. For each cohort, GWAS was performed for each of the 68 iQTs, with global measure of SA or aTH included as covariates. The principal meta-analysis incorporated GWAS results from 33 992 participants, including 23 909 from 49 cohorts involved in ENIGMA consortium and 10 083 from the UK Biobank (http://www.ukbiobank.ac.uk/resources/) [20]. The cortical GWAS summary statistics were acquired from the ENIGMA consortium, encompassing a total of 34 GWAS of regional SA and 34 GWAS of regional aTH.

Given that part of the brain derived QTLs and weights data was specific to the PFC region, our investigation will focus on identifying functional multi-omic factors specific to this area, rather than the entire brain. To this end, we diligently handpicked 10 ROIs located within the PFC region, yielding 20 iQT GWAS (out of the original 68) that were selected. The detailed list of these selected iQTs can be found in Supplementary Table 1.

#### GWAS of brain disorders

We used meta-GWAS summary statistics from five brain disorders (AD, Schizophrenia (SCZ), Major depressive disorder (MDD), Bipolar disorder (BP), and Attention-deficit/hyperactivity disorder (ADHD)) to demonstrate the potential of brain tissue-specific causal factors for providing a contextual understanding of complex brain diseases. Specifically, we sourced the AD GWAS summary statistics from the International Genomics of Alzheimer’s Project (IGAP) GWAS Stage 1 (*N* = 21 982 cases, 41 944 controls), conducted by Kunkle et al. [21]. The GWAS results of SCZ were reported by Trubetskoy et al. [22], with a sample size of 53 386 cases and 77 258 controls. The MDD GWAS summary statistics (*N* = 53 386 cases and 77 258 controls) were acquired from Howard et al. [23]. We obtained the GWAS summary statistics of BP (*N* = 41 917 cases, 371 549 controls) from Mullins et al. [24]. The ADHD GWAS statistics were from Demontis et al. [25] with a sample size of 20 183 cases and 35 191 controls.

### Statistical analysis

#### Transcriptome- and proteome-wide association study

We applied the FUSION software to conduct TWAS and PWAS for each iQT to estimate the associations of each gene and each protein to each brain phenotype. Specifically, the summary-based TWAS model was applied to compute gene-iQT association as the weighted linear sum of SNP-iQT standardized effects (i.e., iQT meta-GWAS z-scores) with the predicted gene-brain effects (i.e., neocortical gene expression weights). Similarly, PWAS was performed by computing the weighted sum of SNP-iQT standardized effects with the pre-computed protein-brain effects (i.e., DLPFC protein abundance weights). We used a threshold of *P* < 1E-3 for both TWAS and PWAS results to define significance. Hereafter, the joint analysis of TWAS and PWAS that identify overlapping findings will be referred to as “TPWAS” for convenience.

#### Transcriptome- and proteome-wide Mendelian randomization analysis

We conducted summary-based transcriptome- and proteome-wide Mendelian randomization (TWMR/PWMR) analysis using the two-sample MR method implemented in the “TwoSampleMR” R package (version 0.5.6, available at <u>mrcieu.github.io/TwoSampleMR</u>). MR analysis investigates the causal relationship between an exposure *X* and an outcome *Y*, using genetic variants as instrumental variables (IVs) *z*. Leveraging the effect sizes of the IVs on both *X* (*β_ZX_*) and *Y* (*β_ZY_*), the MR framework applies the Wald estimate to compute the causal effect of *X* on *Y*: *β_X̂Y_* = *β_zY_*/*β_ZX_*. In a two-sample MR, it is important to acknowledge that statistics may be derived from distinct sample sets. In this study, we integrated the cortical meta-eQTL and DLPFC pQTL data with large-scale iQT GWAS results, to pinpoint risk genes and proteins influencing brain structural alterations. Additionally, we conducted multiple comparison corrections to control for the FDR. Hereafter, the joint analysis of TWMR and PWMR will be referred to as “TPWMR” for convenience.

#### Gene-based analysis of iQT GWAS

To demonstrate the detection capabilities of TPWAS and TPWMR, we compared our findings to the gene-based outcomes derived from applying MAGMA [26] to each of the 20 iQT GWAS datasets. More specifically, we utilized the European samples from the 1 000 Genomes dataset as an LD reference and deployed the SNP-wise mean model to test of the mean SNP associations. A relaxed significance threshold was set at *P* < 1E-4 for fair comparisons.

#### Bayesian colocalization analysis

Bayesian colocalization [27] leverages Bayesian inference to assess the presence of shared genetic variants across two or more traits. It involves analyzing the posterior probability that a variant exhibiting individual associations with each trait or demonstrating a joint association, thereby qualifying as a colocalizing variant. Our analysis incorporated this approach to assess the shared genetic architecture between gene expression and protein abundance in the brain, employing eQTLs and pQTLs extracted from identified genes and proteins. We used the R “coloc” package (https://cran.r-project.org/web/packages/coloc) for testing colocalization, adopting default prior probabilities (i.e., *P*_1_ = 1 × 10^-4^, *P*_2_ = 1 × 10^-4^, and *P*_l2_ = 1 × 10^-5^). Here *P*_l_ and *P*_2_ present the probabilities of a specific variant being associated with gene expression and protein expression, respectively, while *P*_l2_ indicates the likelihood of a given variant simultaneously influencing both gene and protein expressions. A total of five hypotheses (H0∼H4) were tested, with H4 representing the existence of a single causal variant for both traits. We characterized the traits as colocalized when the posterior probability for H4 exceeded 0.8 (i.e., PPH4 > 0.8) [28,29].

#### Cell-type specificity analysis

The cell type-specific analysis of identified genes was further expanded, focusing on their expression levels in human brain cells. For this purpose, we used single-cell RNA-Seq data from the PFC, as profiled by Mathys et al. [30]. We retrieved the raw data and applied the Seurat package for quality control (QC), normalization, and scaling. To be more specific, the initial dataset contained 70 634 single-nucleus transcriptomes. This data was QCed by filtering cells that have unique feature counts over 2 500 or less than 200, as well as genes with fewer than 3 counts in a cell. Global-scaling normalization method “LogNormalize” was employed to normalize the feature expression measurements for each cell by the total expression, and then a linear transformation was used to scale the expression of each gene such that the mean expression across cells for each gene is 0 and the variance is 1. After pre-processing, the cleaned data comprised 17 775 genes across 53 083 cells. We focused on five main cell types: Excitatory neurons, Inhibitory neurons, Oligodendrocytes, Microglia, and Astrocytes. Subsequently, we performed differential expression (DE) analysis for the candidate genes to test if they exhibit significantly higher expressions in certain cell types relative to others. A fold change threshold of 0.25 (i.e., logFC > 0.25) and the Bonferroni corrected *P*-value of 0.05 (adjusted for 17 775 genes) were applied.

#### Causal relationships between brain targets and brain disorders

Prioritizing brain region-specific causal factors could offer insightful perspectives into the complex pathophysiology underlying complex brain disorders, potentially informing more effective therapeutic strategies. Therefore, we further evaluated the potential causal relationships of brain PFC-specific targets on five different brain disorders by performing both TPWAS and TPWMR analyses using GWAS of brain disorders and brain-derived QTLs and weights data. We additionally applied Bonferroni correction to each type of analysis for controlling the multiple comparisons.

## Results

We conducted the brain tissue-specific TPWAS and TPWMR to pinpoint functional elements influencing brain regions at both gene and protein expression levels. We then evaluated the identified genes and proteins through colocalization analysis and cell-type-specific enrichment. To validate our findings, we further assessed their relevance to five brain disorders including AD, SCZ, BP, MDD, and ADHD.

### TPWAS identifies 16 targets associated with PFC iQTs

We utilized meta-GWAS summary statistics of 20 iQTs and employed the summary-based TWAS model trained on neocortical expression data, to estimate the effects of gene expression on PFC ROIs. For each brain iQT, we examined a total of 6 780 genes to determine their associations with the corresponding brain ROIs. Similarly, we applied the PWAS model trained on DLPFC to explore the effects of 1 761 proteins on 20 iQTs.

TWAS and PWAS identified 18 gene-iQT and 8 protein-iQT associations at a false discovery rate (FDR) of 0.05, respectively. In our endeavor to simultaneously explore biomolecules affecting brain regions from both gene expression and protein expression perspectives, we adopted a relatively lenient threshold for circumventing the potential omission of biologically meaningful findings. Using a significance threshold of *P* < 1E-3, the TWAS detected a total of 415 gene-iQT associations, which included 356 genes and 20 iQTs. Simultaneously, the PWAS identified 94 protein-iQT associations between 78 proteins and 20 iQTs. Upon integrating the TWAS and PWAS results, we derived a list of 19 gene/protein-iQT associations between 16 genes/proteins and 12 iQTs that demonstrated significance in both studies (Table 1).

**Table 1.**
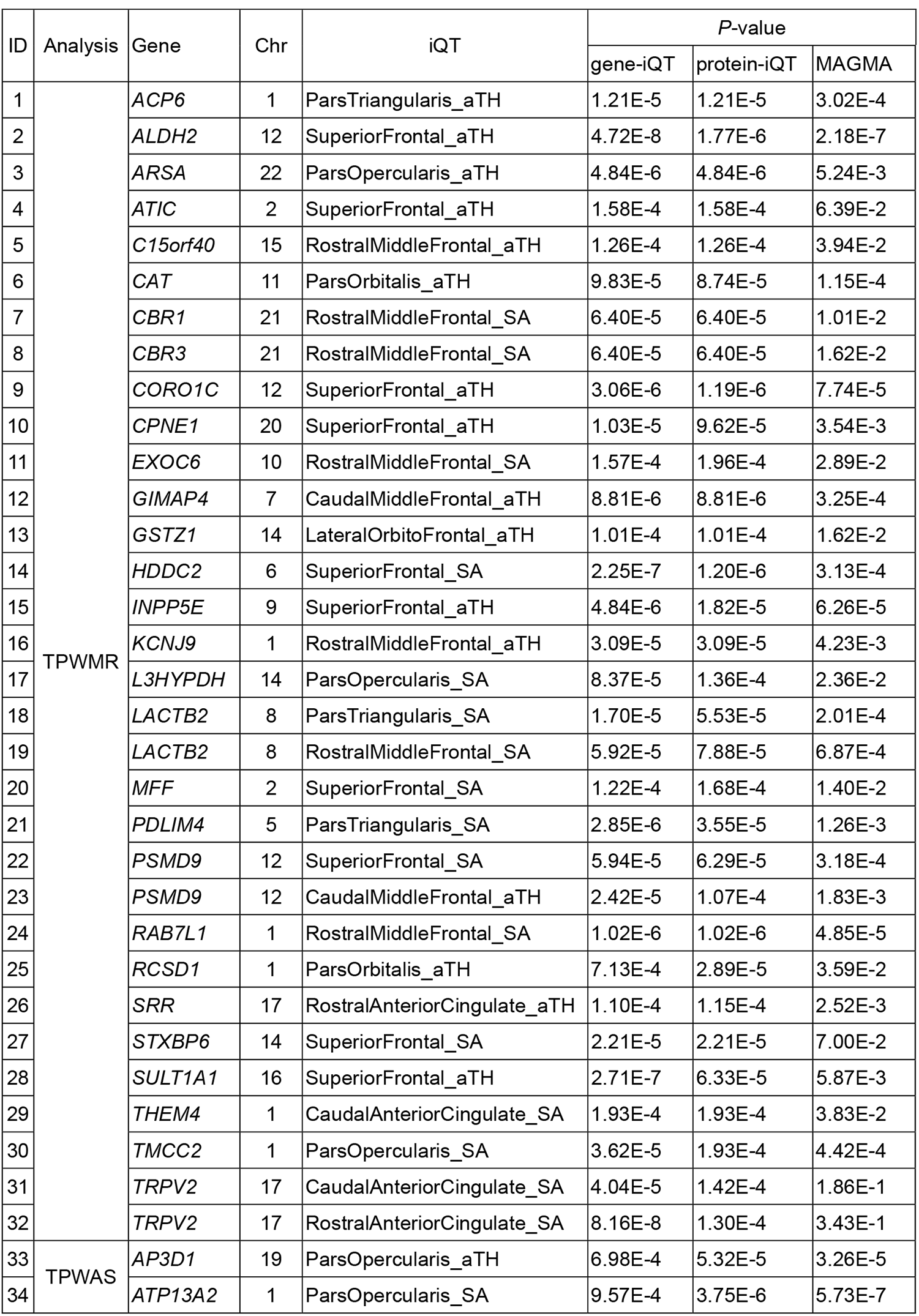
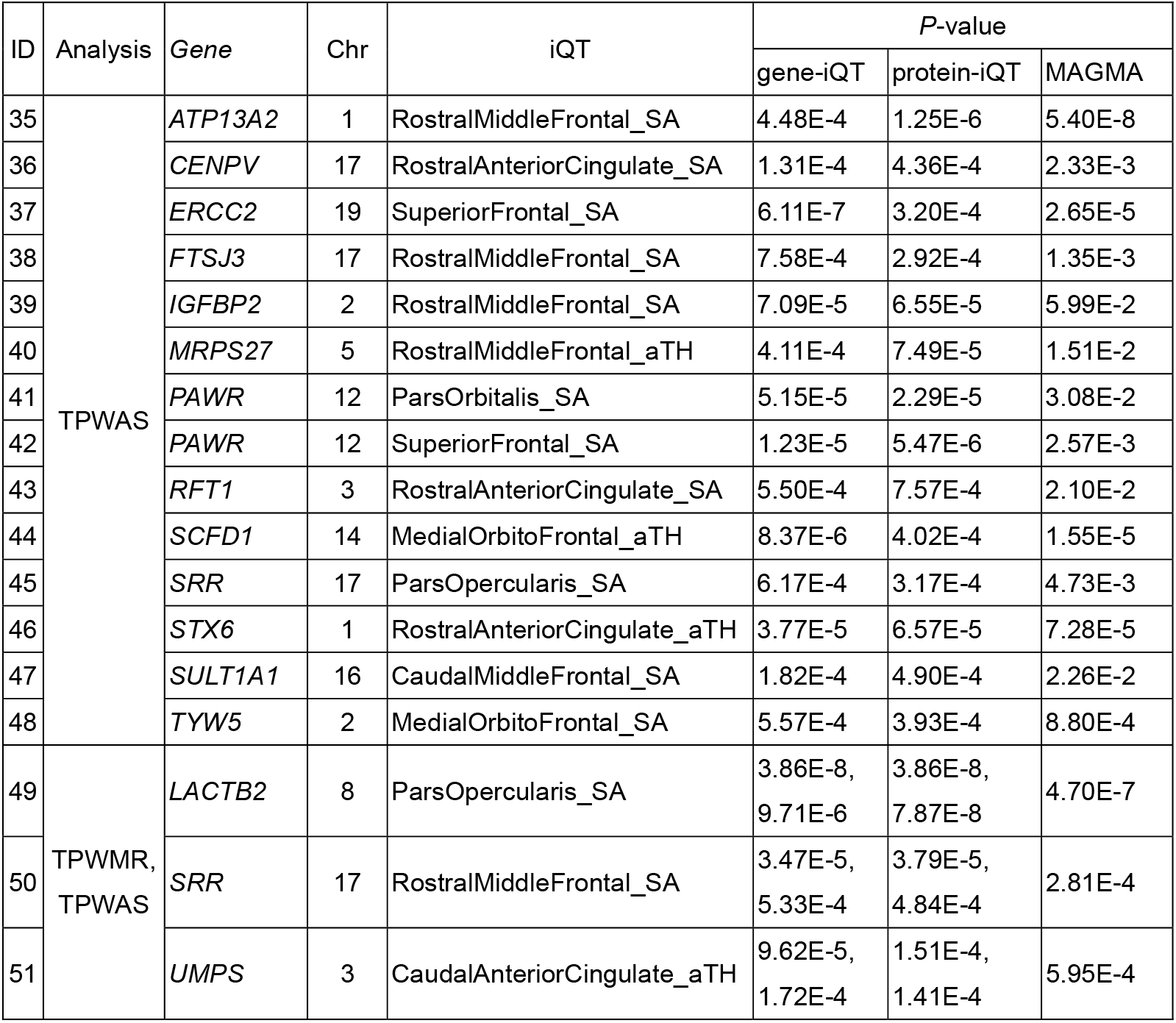
TPWMR and TPWAS identified a total of 51 gene/protein-iQT associations. This led to the discovery of 42 distinct targets associated with 19 iQTs across 10 prefrontal cortical regions. 11 out of the 51 findings were detected by the MAGMA with a significant *P* < 1E-4. Detailed information is available in Supplementary Table 1.

### TPWMR identifies 30 targets associated with PFC ROIs

In our summary-based MR analysis, we integrated meta-GWAS data from 20 iQTs with brain cortical eQTLs and DLPFC-derived pQTLs, respectively, aiming to discover genes and proteins whose expression alterations impact brain structures through eQTL or pQTL SNPs. Particularly, the Wald ratio was applied for proposed instruments with one SNP and the inverse variance weighted (IVW) test was employed for proposed instruments with more than one SNP. TWMR and PWMR identified 3 642 gene-iQT and 61 protein-iQT associations at FDR < 0.01, respectively. Upon integrating TWMR and PWMR, we compiled a list of 35 gene/protein-iQT associations between 30 genes/proteins and 15 iQTs that demonstrated significance in TPWMR (Table 1).

### Joint TPWAS-TPWMR identifies 42 targets associated with 19 PFC ROIs

We obtained 51 gene/protein-iQT associations by integrating findings from TPWAS and TPWMR. Table 1 and Supplementary Table 2 illustrate the details of identified gene/protein-iQT associations. This comprehensive analysis led to the discovery of 42 distinct genes associated with 10 ROIs across 19 iQTs. Figure 2 depicted the pairwised associations between all of the 42 targets and the 19 iQTs as determined by both the TPWAS and TPWMR methods. Of the 51 significant findings, 1) three (5.8%) associations were reported by both TPWAS and TPWMR analyses, including the association between *LACTB2* with the surface area of pars opercularis, the association between *SRR* with the surface area of the rostral middle frontal lobe, and the association between *UMPS* with the cortical thickness of caudal anterior cingulate; 2) one gene (*SULT1A1*) was identified through two distinct methods, with each method associating with different iQTs. In the TPWMR analysis, *SULT1A1* was found to be associated with the cortical thickness of the superior frontal lobe, while the TPWAS analysis linked *SULT1A1* to the surface area of the caudal middle frontal gyrus; and 3) five genes (12%) demonstrated significant associations with more than one neuroimaging traits in either TPWAS or TPWMR analysis, including *ATP13A2*, *LACTB2*, *PAWR*, *PSMD9*, and *TRPV2*. Amongst the 51 identified associations, we discovered that only 11 findings, accounting for 21.6%, were detected by MAGMA as shown in Table 1.

**Figure. 2.**
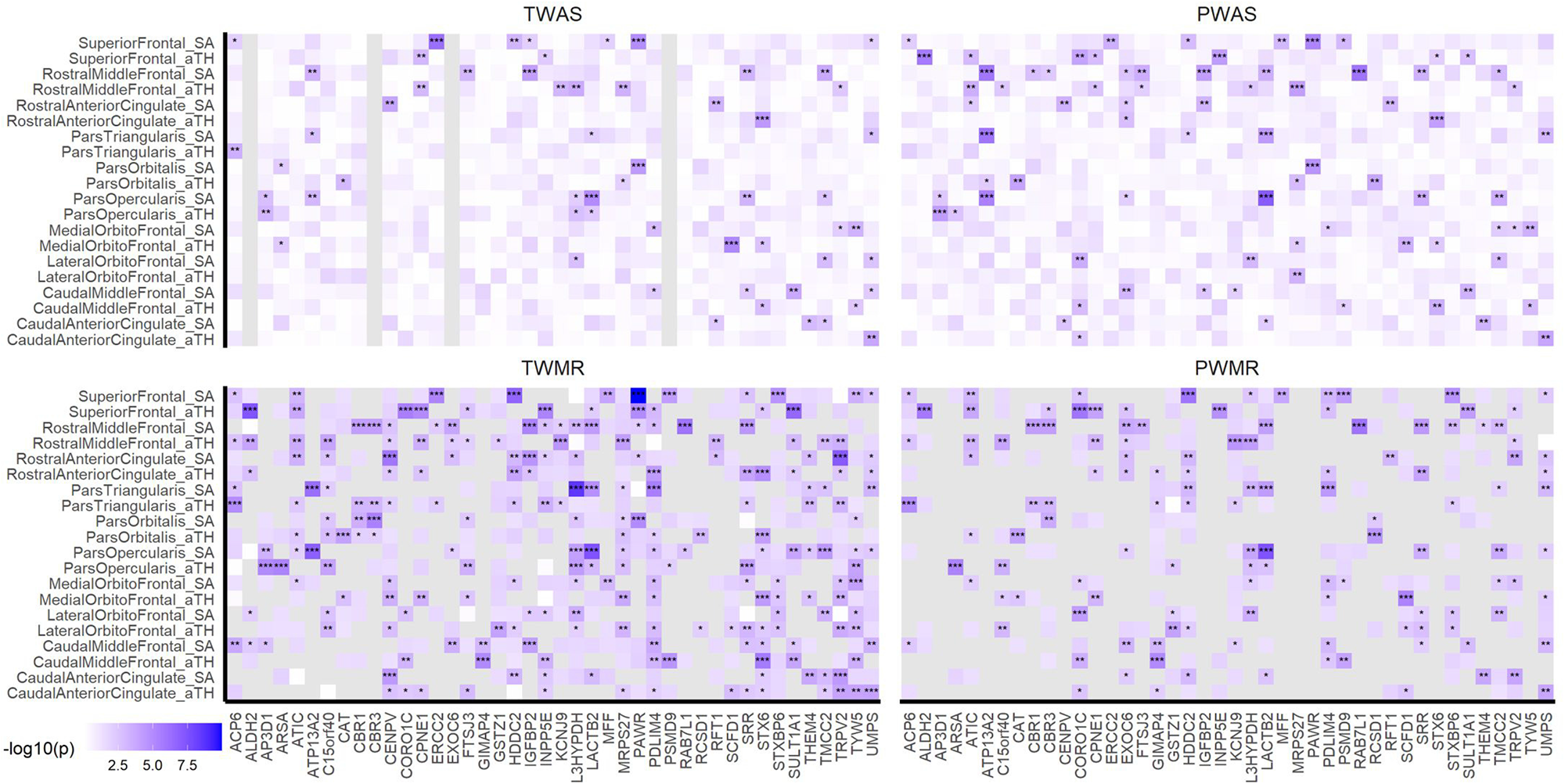
Heatmaps of gene associations on imaging quantitative traits (iQTs). The heatmap was generated by extracting associations between 42 genes and 19 iQTs from the TWAS, PWAS, TWMR and PWMR results. Heatmap cells are marked with “*”, “**” or “***” signifying significance levels of 0.001 < *P* < 0.01, 0.0001 < *P* < 0.001, and *P* < 0.0001, respectively. Cells corresponding to NAN values are shaded gray.

### Colocalization of gene and protein expressions

The colocalization analysis was carried out for 42 identified targets, with the detailed results presented in Table 2. Our study found that 32 (76.2%) genes/proteins exhibited signs of sharing causal variants in their gene and protein expression (i.e., with a PPH4 > 0.8). The Locuszoom plots (http://locuszoom.org/ [31]) highlighted the evidence of colocalization between genetic loci and their corresponding gene and protein expressions. See Supplementary Figure 1 for detailed Locuszoom plots.

**Table 2.** Results of the colocalization analysis for the 42 candidate genes and their corresponding protein expressions. Genes/proteins demonstrating a posterior probability greater than 0.8 for hypothesis 4 (i.e., PPH4 > 0.8) are highlighted in bold.

| Gene | Chr | Start | End | nsnps | PPH0 | PPH1 | PPH2 | PPH3 | PPH4 |
| --- | --- | --- | --- | --- | --- | --- | --- | --- | --- |
| *ACP6* | 1 | 147101453 | 147142618 | 127 | 0.000 | 0.000 | 0.000 | 1.000 | 0.000 |
| ***ALDH2*** | 12 | 112204691 | 112247782 | 61 | 0.000 | 0.000 | 0.000 | 0.027 | **0.973** |
| ***AP3D1*** | 19 | 2100988 | 2164464 | 72 | 0.000 | 0.127 | 0.000 | 0.008 | **0.865** |
| ***ARSA*** | 22 | 51061182 | 51066607 | 99 | 0.000 | 0.000 | 0.000 | 0.008 | **0.992** |
| ***ATIC*** | 2 | 216176540 | 216214487 | 130 | 0.000 | 0.000 | 0.000 | 0.000 | **1.000** |
| ***ATP13A2*** | 1 | 17312453 | 17338423 | 113 | 0.000 | 0.023 | 0.000 | 0.004 | **0.973** |
| *C15orf40* | 15 | 83657193 | 83680393 | 69 | 0.000 | 0.000 | 0.000 | 1.000 | 0.000 |
| ***CAT*** | 11 | 34460472 | 34493609 | 188 | 0.000 | 0.000 | 0.000 | 0.006 | **0.994** |
| ***CBR1*** | 21 | 37442239 | 37445464 | 115 | 0.000 | 0.000 | 0.000 | 0.034 | **0.966** |
| ***CBR3*** | 21 | 37507210 | 37518864 | 122 | 0.000 | 0.000 | 0.000 | 0.003 | **0.997** |
| ***CENPV*** | 17 | 16245848 | 16256970 | 73 | 0.000 | 0.000 | 0.000 | 0.007 | **0.993** |
| ***CORO1C*** | 12 | 109038885 | 109125372 | 121 | 0.000 | 0.000 | 0.000 | 0.010 | **0.990** |
| ***CPNE1*** | 20 | 34213953 | 34252878 | 84 | 0.000 | 0.000 | 0.000 | 0.002 | **0.998** |
| *ERCC2* | 19 | 45853095 | 45874176 | 63 | 0.000 | 0.612 | 0.000 | 0.149 | 0.239 |
| ***EXOC6*** | 10 | 94590935 | 94819250 | 218 | 0.000 | 0.000 | 0.000 | 0.019 | **0.981** |
| ***FTSJ3*** | 17 | 61896793 | 61907372 | 68 | 0.000 | 0.000 | 0.000 | 0.039 | **0.961** |
| *GIMAP4* | 7 | 150264365 | 150271041 | 114 | 0.000 | 0.000 | 0.000 | 0.997 | 0.003 |
| ***GSTZ1*** | 14 | 77787227 | 77797940 | 128 | 0.000 | 0.000 | 0.000 | 0.073 | **0.927** |
| ***HDDC2*** | 6 | 125541108 | 125623282 | 135 | 0.000 | 0.000 | 0.000 | 0.002 | **0.998** |
| ***IGFBP2*** | 2 | 217497551 | 217529159 | 119 | 0.000 | 0.006 | 0.000 | 0.002 | **0.992** |
| *INPP5E* | 9 | 139323071 | 139334274 | 107 | 0.000 | 0.009 | 0.000 | 0.980 | 0.011 |
| ***KCNJ9*** | 1 | 160051360 | 160060353 | 139 | 0.000 | 0.000 | 0.000 | 0.145 | **0.855** |
| ***L3HYPDH*** | 14 | 59927081 | 59951148 | 111 | 0.000 | 0.000 | 0.000 | 0.025 | **0.975** |
| *LACTB2* | 8 | 71547553 | 71581409 | 67 | 0.000 | 0.000 | 0.000 | 0.887 | 0.113 |
| *MFF* | 2 | 228189867 | 228222550 | 108 | 0.000 | 0.000 | 0.000 | 0.999 | 0.001 |
| **MRPS27** | 5 | 71515236 | 71616473 | 113 | 0.000 | 0.017 | 0.000 | 0.004 | **0.979** |
| **PAWR** | 12 | 79968759 | 80084877 | 65 | 0.000 | 0.101 | 0.000 | 0.040 | **0.858** |
| **PDLIM4** | 5 | 131593364 | 131609147 | 123 | 0.000 | 0.000 | 0.000 | 0.000 | **1.000** |
| **PSMD9** | 12 | 122326637 | 122356203 | 89 | 0.000 | 0.000 | 0.000 | 0.011 | **0.989** |
| **RAB7L1** | 1 | 205737114 | 205744588 | 92 | 0.000 | 0.000 | 0.000 | 0.001 | **0.998** |
| *RCSD1* | 1 | 167599330 | 167675486 | 197 | 0.000 | 0.000 | 0.155 | 0.184 | 0.661 |
| **RFT1** | 3 | 53122499 | 53164478 | 88 | 0.000 | 0.000 | 0.000 | 0.006 | **0.994** |
| **SCFD1** | 14 | 31091318 | 31205018 | 136 | 0.000 | 0.000 | 0.000 | 0.010 | **0.990** |
| **SRR** | 17 | 2206677 | 2228554 | 112 | 0.000 | 0.000 | 0.000 | 0.000 | **1.000** |
| **STX6** | 1 | 180941861 | 180992047 | 115 | 0.000 | 0.001 | 0.000 | 0.007 | **0.992** |
| **STXBP6** | 14 | 25278862 | 25519503 | 283 | 0.000 | 0.000 | 0.000 | 0.000 | **1.000** |
| **SULT1A1** | 16 | 28616903 | 28634946 | 44 | 0.000 | 0.000 | 0.000 | 0.077 | **0.923** |
| *THEM4* | 1 | 151846060 | 151882284 | 104 | 0.000 | 0.000 | 0.000 | 1.000 | 0.000 |
| **TMCC2** | 1 | 205197304 | 205242471 | 121 | 0.000 | 0.000 | 0.000 | 0.027 | **0.972** |
| **TRPV2** | 17 | 16318856 | 16340317 | 82 | 0.000 | 0.000 | 0.000 | 0.001 | **0.999** |
| *TYW5* | 2 | 200794698 | 200820459 | 74 | 0.000 | 0.232 | 0.000 | 0.044 | 0.724 |
| **UMPS** | 3 | 124449213 | 124464040 | 145 | 0.000 | 0.000 | 0.000 | 0.005 | **0.995** |

### Cell-type-specific enrichment in PFC

We examined the cell-type specificity of reported genes by incorporating human brain single-cell RNA-Seq data. Of the 42 genes considered, the dataset contained information for 19. Our study uncovered over-expression patterns of 12 causal genes in various types of brain cells, ascertained at a Bonferroni corrected *P* < 0.05 and logFC > 0.25. Specifically, the genes *ACP6*, *ALDH2*, *CAT* and *HDDC2* were found to be enriched in astrocytes, while *RCSD1* was enriched in microglia. In oligodendrocytes, four genes (*CBR1*, *CORO1C*, *STXBP6* and *TMCC2*) were observed to be overexpressed, among which *STXBP6* was also found to be enriched in inhibitory neurons. *ATP13A2*, *EXOC6* and *KCNJ9* were found to be enriched in excitatory neurons, whereas *ATP13A2* also showed enrichment in inhibitory neurons. Detailed information was shown in Figure 3 and Supplementary Table 3.

**Figure. 3.**
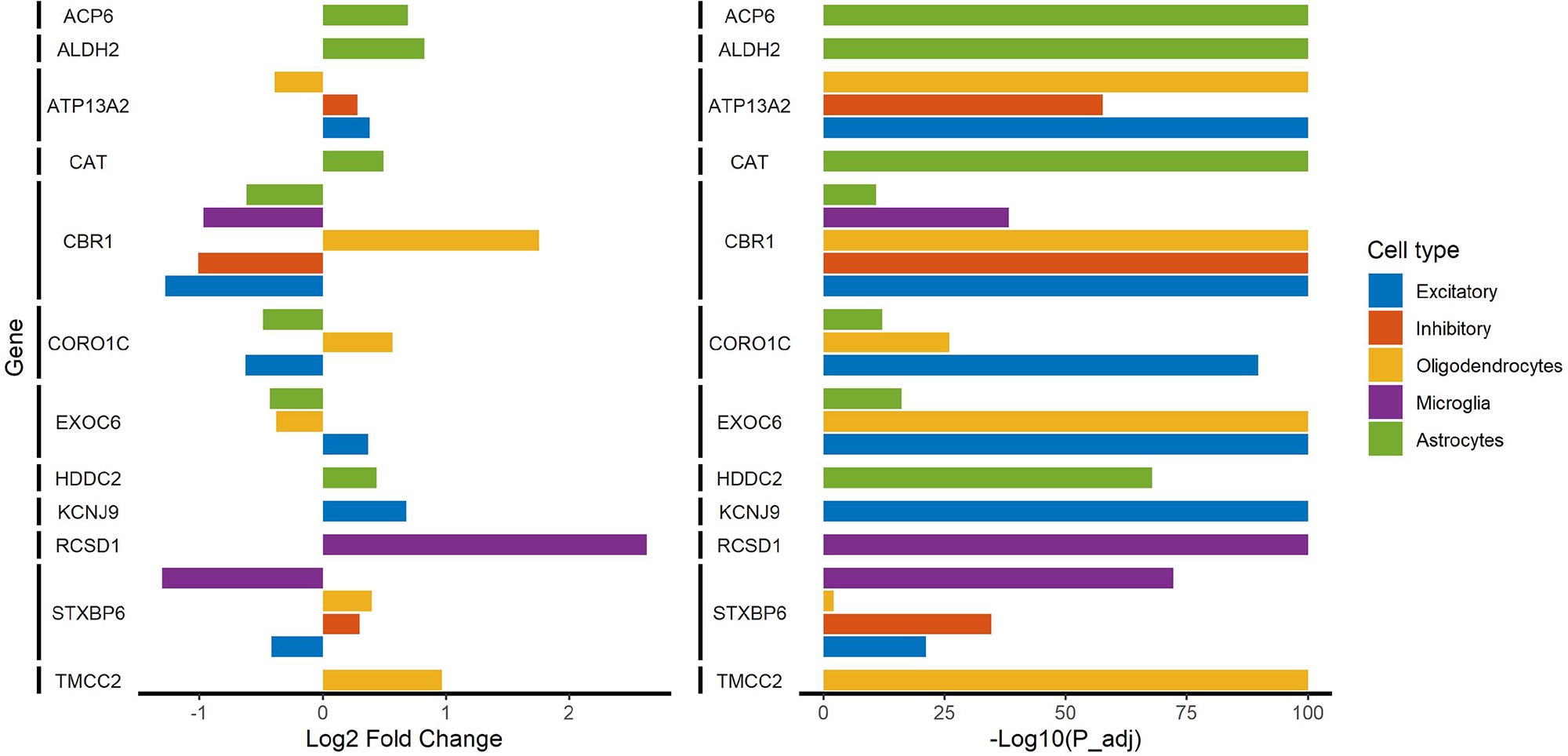
Prefrontal cortex cell-type-specific enrichment of causal genes. Single-cell expression enrichment identified over-expression of 12 causal genes in brain cell types. Left and right panels showed fold change and *P*-value respectively.

### Associations of iQT targets with brain disorders

The exploration of brain region-specific causal factors holds considerable promise in providing a nuanced comprehension of intricate neurological diseases. Our investigation yielded a total of 38 disease-gene/protein associations involving 20 unique molecular entities across all five brain disorders, as detailed in Table 3. Among these significant associations we identified, three were detected via TWAS or PWAS, while the remaining 35 were detected by the MR method. More specifically, 1) we validated varying numbers of targets for each brain disorder: 6 for AD, 6 for ADHD, 7 for BP, 4 for MDD, and 8 for SCZ; 2) five genes (*ARSA*, *GSTZ1*, *L3HYPDH*, *RFT1* and *TYW5*) were found to be associated with more than one brain disease; 3) four genes were each associated with a singular disorder yet were supported by more than one analytical methods, including *ACP6*-MDD, *C15orf40*-BP, *CBR3*-SCZ and *PDLIM4*-MDD associations; and 4) three genes (*RAB7L1*, *SCFD1* and *SRR*) demonstrated associations with multiple brain diseases and were additionally corroborated by more than one type of analysis. Summarized results and detailed information regarding comparison correction were presented in Supplementary Table 4 and Supplementary Table 5.

**Table 3.** Validation of associations between 20 iQT targets with five brain disorders.

| Brain disorder | Analysis | Gene | Chr | Start | End | z | P-value |
| --- | --- | --- | --- | --- | --- | --- | --- |
| AD | TWMR | *FTSJ3* | 17 | 61896793 | 61907372 | 4.097 | 4.19E-05 |
|  | TWMR | *MFF* | 2 | 228189867 | 228222550 | -3.626 | 2.88E-04 |
|  | TWMR | *MRPS27* | 5 | 71515236 | 71616473 | -3.591 | 3.30E-04 |
|  | TWMR | *SRR* | 17 | 2206677 | 2228554 | 3.958 | 7.54E-05 |
|  | TWMR | *STX6* | 1 | 180941861 | 180992047 | 3.497 | 4.71E-04 |
|  | TWMR | *TYW5* | 2 | 200794698 | 200820459 | -4.497 | 6.88E-06 |
| ADHD | TWMR | *ALDH2* | 12 | 112204691 | 112247782 | -4.684 | 2.81E-06 |
|  | PWMR | *GSTZ1* | 14 | 77787227 | 77797940 | -4.102 | 4.09E-05 |
|  | TWMR | *L3HYPDH* | 14 | 59927081 | 59951148 | -3.799 | 1.45E-04 |
|  | PWMR | *SCFD1* | 14 | 31091318 | 31205018 | -3.727 | 1.94E-04 |
|  | TWMR | *SRR* | 17 | 2206677 | 2228554 | -4.585 | 4.53E-06 |
|  | TWMR | *SULT1A1* | 16 | 28616903 | 28634946 | -4.614 | 3.94E-06 |
| BP | TWMR | *ARSA* | 22 | 51061182 | 51066607 | 4.306 | 1.66E-05 |
|  | PWMR | *C15orf40* | 15 | 83657193 | 83680393 | 5.522 | 3.35E-08 |
|  | TWAS | *C15orf40* | 15 | 83657193 | 83680393 | 4.984 | 6.24E-07 |
|  | TWMR | *GSTZ1* | 14 | 77787227 | 77797940 | -3.734 | 1.89E-04 |
|  | PWMR | *L3HYPDH* | 14 | 59927081 | 59951148 | 3.528 | 4.18E-04 |
|  | PWAS | *RAB7L1* | 1 | 205737114 | 205744588 | 3.841 | 1.23E-04 |
|  | PWMR | *RAB7L1* | 1 | 205737114 | 205744588 | 3.841 | 1.23E-04 |
|  | PWMR | *RFT1* | 3 | 53122499 | 53164478 | 3.408 | 6.55E-04 |
|  | PWMR | *SRR* | 17 | 2206677 | 2228554 | -4.094 | 4.24E-05 |
|  | TWMR | *SRR* | 17 | 2206677 | 2228554 | -3.891 | 9.99E-05 |
| MDD | PWMR | *ACP6* | 1 | 147101453 | 147142618 | -3.744 | 1.81E-04 |
|  | TWMR | *ACP6* | 1 | 147101453 | 147142618 | -3.744 | 1.81E-04 |
|  | PWMR | *PDLIM4* | 5 | 131593364 | 131609147 | -3.790 | 1.50E-04 |
|  | TWMR | *PDLIM4* | 5 | 131593364 | 131609147 | -3.984 | 6.78E-05 |
|  | TWMR | *RAB7L1* | 1 | 205737114 | 205744588 | 4.227 | 2.37E-05 |
|  | TWMR | *RFT1* | 3 | 53122499 | 53164478 | 4.327 | 1.51E-05 |
| SCZ | TWMR | *AP3D1* | 19 | 2100988 | 2164464 | 3.977 | 6.99E-05 |
|  | TWMR | *ARSA* | 22 | 51061182 | 51066607 | 5.374 | 7.71E-08 |
|  | PWMR | *CBR3* | 21 | 37507210 | 37518864 | -4.330 | 1.49E-05 |
|  | TWMR | *CBR3* | 21 | 37507210 | 37518864 | -3.885 | 1.02E-04 |
|  | PWMR | *L3HYPDH* | 14 | 59927081 | 59951148 | 3.378 | 7.30E-04 |
|  | PWMR | *SCFD1* | 14 | 31091318 | 31205018 | -3.708 | 2.09E-04 |
|  | TWMR | *SCFD1* | 14 | 31091318 | 31205018 | -3.708 | 2.09E-04 |
|  | PWMR | *SRR* | 17 | 2206677 | 2228554 | -5.348 | 8.91E-08 |
|  | TWAS | *TYW5* | 2 | 200794698 | 200820459 | -4.476 | 7.60E-06 |
|  | PWMR | *UMPS* | 3 | 124449213 | 124464040 | -3.413 | 6.42E-04 |

## Discussion

Emphasizing the role of brain region-specific factors may afford substantial insights into the intricate pathophysiological underpinnings of complex brain disorders, which could subsequently guide the development of more effective therapeutic interventions. In this study, we systematically integrated brain tissue-specific multi-omics data by comprehensively applying TWAS, PWAS, MR, and Bayesian colocalization. In summary, we discovered 51 gene-iQT associations that linked 42 targets with 10 brain prefrontal cortical regions, at both transcriptome and proteome levels. Colocalization between gene and protein expressions pointed towards a significant shared genetic underpinning between gene transcription and protein synthesis in the brain, thereby unveiling a substantial biological interdependence.

Of the 51 identified associations, 40 gene-iQT relationships were not discovered by the GWAS of the same ENIGMA study. This suggests that exclusively relying on GWAS or post-GWAS screening may not be sufficient for effectively identifying genes of relevance to brain functions or disorders. This highlights the necessity for diversification and supplementation of methods in gene identification processes.

The validation of the causal effects of 20 (out of 42) brain targets on complex brain disorders emphasizes the potential of investigating brain structural alterations in the study of neuropsychiatric diseases. Future study could explore the relationships among these identified genes/proteins, brain regional structures and various brain disorders, for example, the mediating role of brain regions between gene/protein expression and brain diseases. Meanwhile, for the 22 targets showing no explicit disease relevance, further investigation into their regulatory mechanisms influencing brain changes could be valuable. This is especially important considering the extensive pathways from gene to cognitive conditions, as well as the complex mechanisms intertwining brain alteration and disease development.

Our analysis implicates a set of genes that have been previously investigated in relation to diverse brain phenotypes and brain disorders. For example, *ACP6*, a lysophosphatidic acid phosphatase that regulates mitochondrial lipid biosynthesis, is associated with cerebral impairment-induced visual disorder [32], Parkinson’s disease (PD) [33] and autism spectrum disorders [34]. *ALDH2* is one of the major ALDH isozymes that catalyze the oxidation of dopamine. Impairment or inactivation of *ALDH2* would accumulate a higher level of toxic dopamine metabolite, consequently contributing to neurocognitive dysfunction [35,36]. Brain atrophy and AD biomarkers changes were also observed from *Aldh2-/-* mice model [37]. *SRR* has been reported to be the principal enzyme responsible for D-serine production in the mouse forebrain, the latter plays an important role in mammalian brain neurotransmission [38]. *RAB7L1*, the second PD-associated gene, has been linked to the induction of neuronal apoptosis and damage, as well as learning and cognitive dysfunctions, which highlights its possible implication in neurodegenerative and cognitive disorders [39–41]. Dysregulation or loss of *ATP13A2* function can affect numerous regions within the nervous system, leading to a wide range of symptoms, including dementia, spasticity, and parkinsonism [42,43]. Additionally, *ATP13A2* presents reduced protein levels in brain tissue of cases with Lewy bodies [42]. This convergence of findings reinforces the potential role of these genes in the pathophysiology of neurological impairments and provides an expanded platform for further research and therapeutic development. We also suggest the potential roles of candidate findings in various cell types, some of which have been previously explored. For example, the expression of *CAT* in astrocytes may play a role in protecting the brain from oxidative stress and damage [44].

Linking the brain tissue-specific causal factors to brain disorders holds considerable promise in providing a nuanced comprehension of intricate neurological diseases. Our explorations implicate a set of genes for their possible broad-spectrum involvement in neural pathology. Moreover, the multi-method support further strengthens the likelihood of some identified associations. Collectively, these validations underscore the complexity of gene-disease associations in brain disorders and the utility of multi-omics and systematical approaches in unveiling these relationships. Recognizing the unique contributions of distinct brain regions to overall neural function, it is reasonable to hypothesize that the pathogenesis of neurophysiological disorders may be tightly linked with dysfunctions or dysregulations localized to specific regions. Therefore, a contextual approach that prioritizes region-specific etiological factors could offer insightful perspectives into the complex pathophysiology underlying these disorders, potentially informing more effective therapeutic strategies.

In conclusion, we provide a novel sight into integrating multi-omics data to explore the molecular mechanisms underlying the brain. Neuroimaging serves as a pivotal biomarker for early diagnosis of brain disorders, thereby rendering the omics research of brain imaging of profound significance. This is particularly relevant given our deployment of various methodologies for the systematic integration of multiple omics data. The advancements in omics technologies and the enrichment of omics data set stage for the future execution of multi-omics inference research across a wider range of brain tissues. This endeavor will require a transition from single-omics to a more comprehensive pan-omics approach, further incorporating dynamic omics studies that span the entirety of the life course. These integrative strategies promise to yield insights into the complex temporal and spatial dynamics of neurological development and function.

## Supporting information

Colocalization between gene and protein expressions

20 iQTs that derived from the prefrontal cortex (PFC) were included in this study

## Data Availability

All data produced in the present work are contained in the manuscript

## DATA AVAILABILITY

The brain eQTL and TWAS weights can be accessed at https://doi.org/10.7303/syn16984815.1 and https://doi.org/10.7303/syn2580853, respectively. The brain pQTL data and PWAS weights from the ROS/MAP study are available at https://doi.org/10.7303/syn23627957. The cortical GWAS summary statistics from the ENIGMA consortium are accessible at https://enigma.ini.usc.edu/research/download-enigma-gwas-results/ upon request.

The GWAS summary statistics of AD are available from NIAGADS under accession NG00075 (https://www.niagads.org/datasets/ng00075), and the GWAS summary statistics of ADHD, BP, MDD, and SCZ can be accessed at https://www.med.unc.edu/pgc/download-results/. The human brain cell-type RNA-seq data from the ROS/MAP study are available at https://doi.org/10.7303/syn18485175.

## CODE AVAILABILITY

The code used throughout this study is available upon reasonable request from the corresponding authors.

## ACKNOWLEDGEMENTS

This work is supported in part by the National Natural Science Foundation of China (62103116, 62102115), the Shandong Provincial Natural Science Foundation (2022HWYQ-093), the Natural Science Foundation of Heilongjiang Province (LH2022F016), and by the Fundamental Research Funds for the Central Universities (3072022TS2614, 79000010, 79000011).

**AMPAD Acknowledgements** The results published here are in whole or in part based on data obtained from the AD Knowledge Portal (https://adknowledgeportal.org).

**IGAP Acknowledgements** We thank the International Genomics of Alzheimer’s Project (IGAP) for making summary results data available to the public, which were used in our analysis. The investigators within IGAP contributed to the design and implementation of IGAP and/or provided data but did not participate in analysis or writing of this report. IGAP was made possible by the generous participation of the control subjects, the patients, and their families. The i–Select chips was funded by the French National Foundation on Alzheimer’s disease and related disorders. EADI was supported by the LABEX (laboratory of excellence program investment for the future) DISTALZ grant, Inserm, Institut Pasteur de Lille, Université de Lille 2 and the Lille University Hospital. GERAD was supported by the Medical Research Council (Grant n° 503480), Alzheimer’s Research UK (Grant n° 503176), the Wellcome Trust (Grant n° 082604/2/07/Z) and German Federal Ministry of Education and Research (BMBF): Competence Network Dementia (CND) grant n° 01GI0102, 01GI0711, 01GI0420. CHARGE was partly supported by the NIH/NIA grant R01 AG033193 and the NIA AG081220 and AGES contract N01–AG–12100, the NHLBI grant R01 HL105756, the Icelandic Heart Association, and the Erasmus Medical Center and Erasmus University. ADGC was supported by the NIH/NIA grants: U01 AG032984, U24 AG021886, U01 AG016976, and the Alzheimer’s Association grant ADGC–10–196728.

