## Supplementary material for "Integration of tissue-specific multi-omics data implicates brain targets for complex neuropsychiatric traits": Colocalization between gene and protein expressions

<u>Supplementary Figures</u>

**Integration of tissue-specific multi-omics data to predict brain biomarkers**

*** Correspondence to:**

**Supplementary Figure 1. Colocalization between gene and protein expressions.** Locuszoom plots illustrate the corresponding eQTLs and pQTLs for each candidate target. The color scale of $r^2$ values is used to label SNPs based on their degree of linkage disequilibrium with the relevant peak eQTL or pQTL.

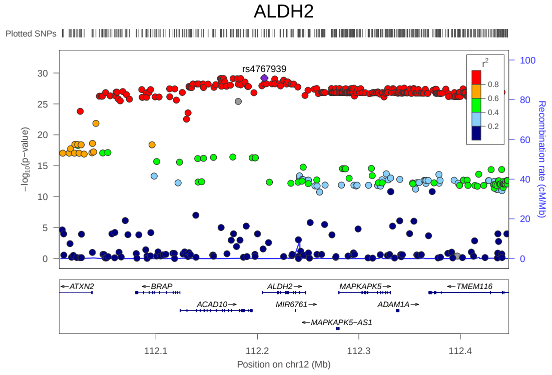


(1) eQTL of ALDH2

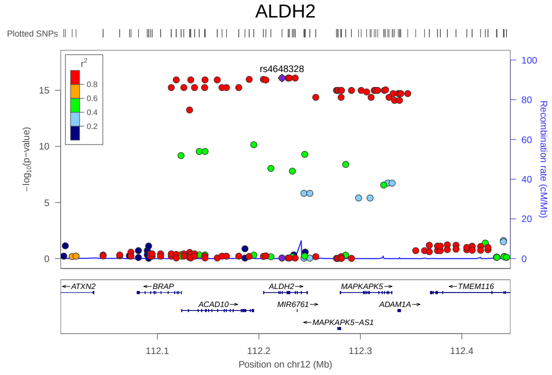


(2) pQTL of ALDH2

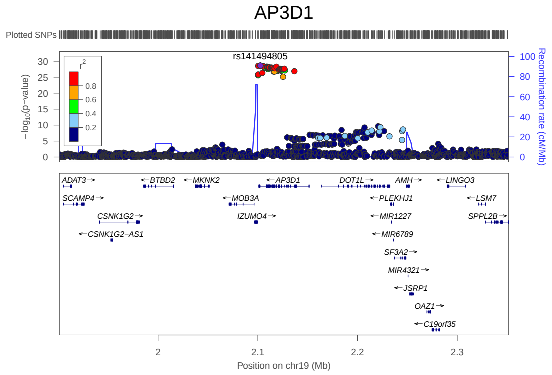


(3) eQTL of AP3D1

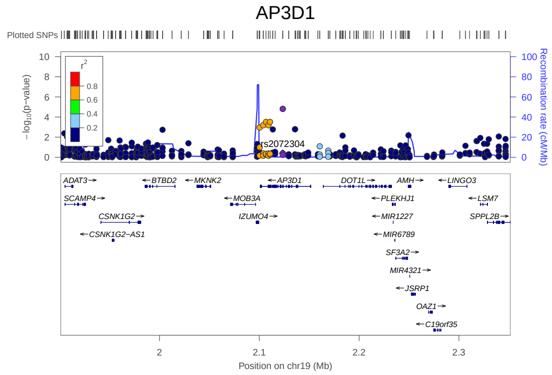


(4) pQTL of AP3D1

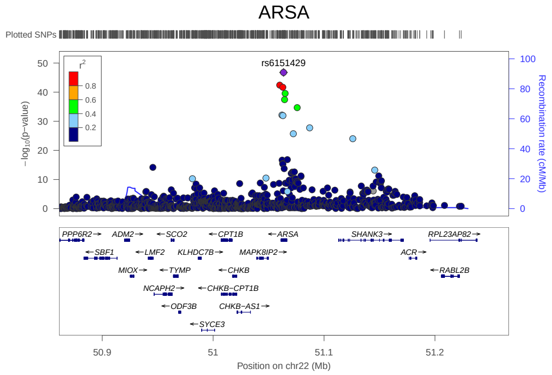


(5) eQTL of ARSA

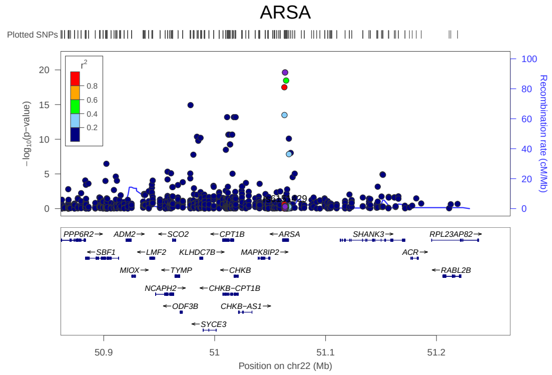


(6) pQTL of ARSA

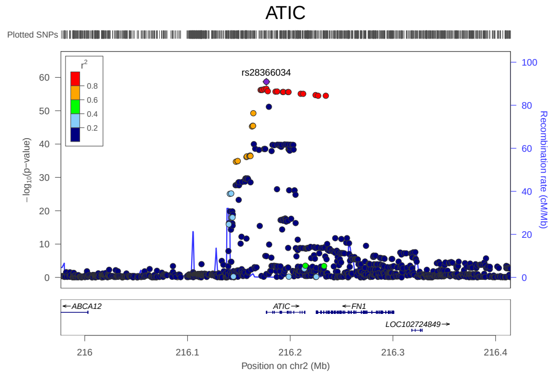


(7) eQTL of ATIC

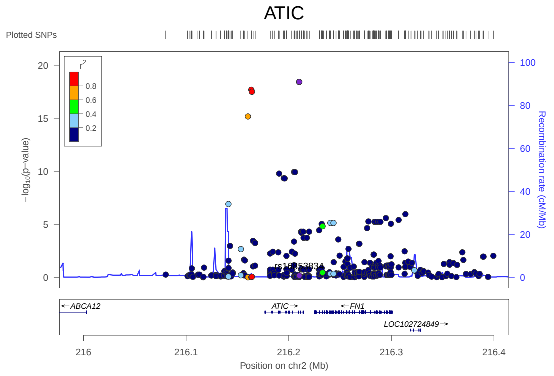


(8) pQTL of ATIC

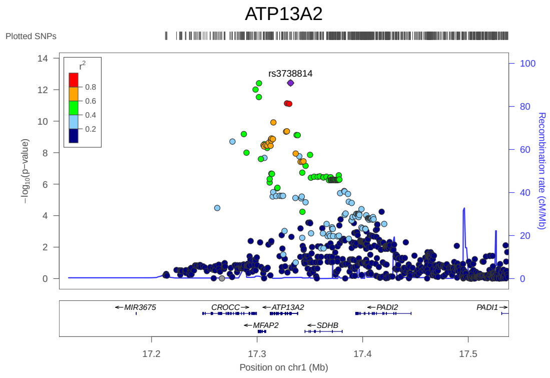


(9) eQTL of ATP13A2

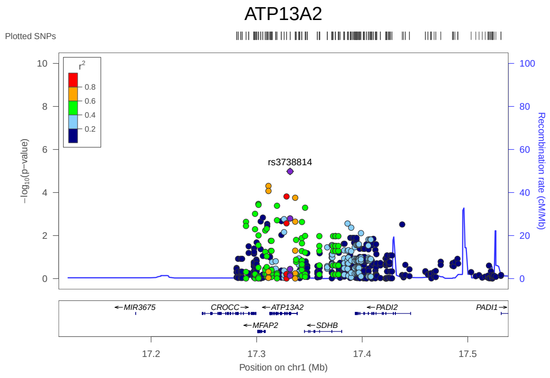


(10) pQTL of ATP13A2

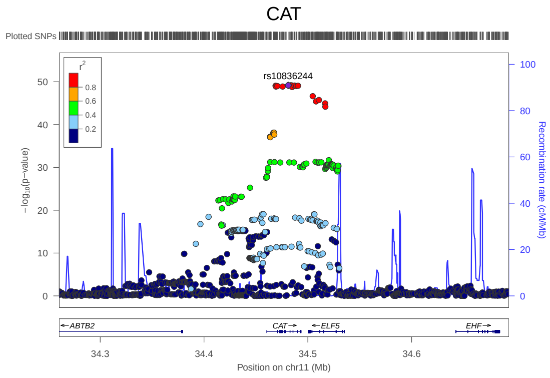


(11) eQTL of CAT

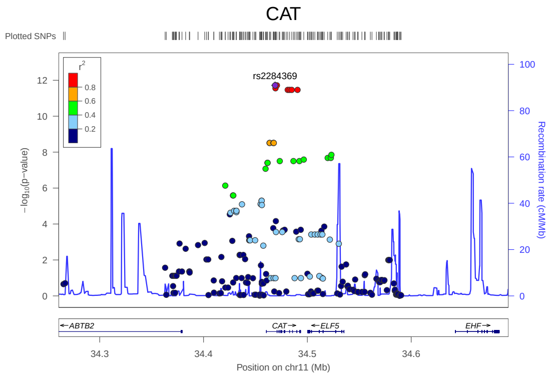


(12) pQTL of CAT

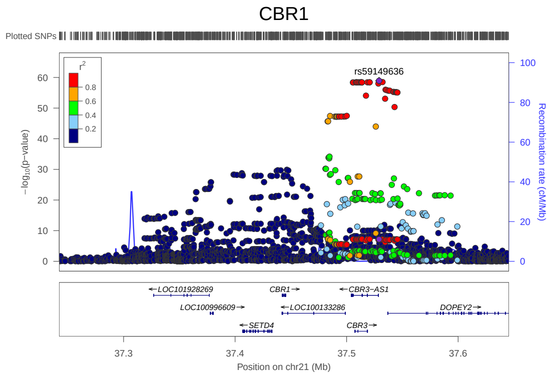


(13) eQTL of CBR1

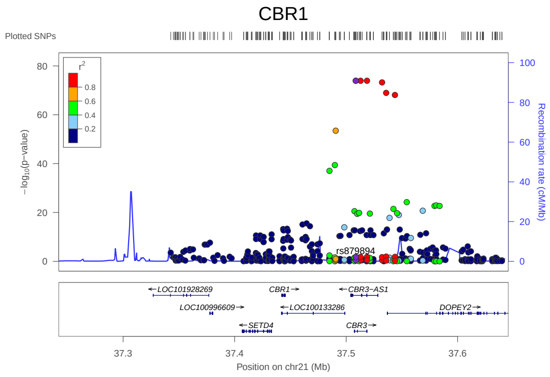


(14) pQTL of CBR1

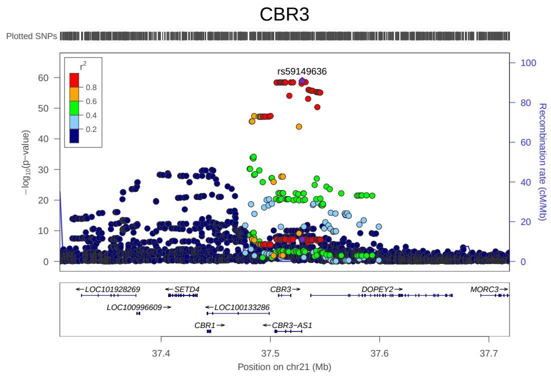


(15) eQTL of CBR3

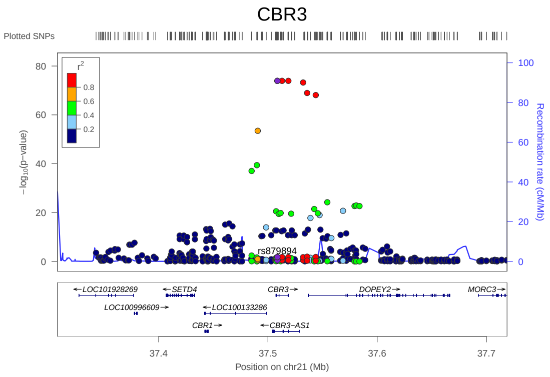


(16) pQTL of CBR3

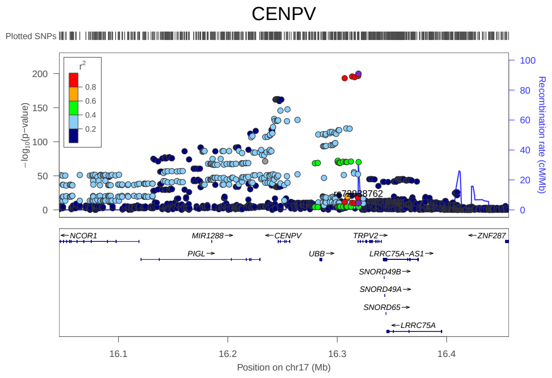


(17) eQTL of CENPV

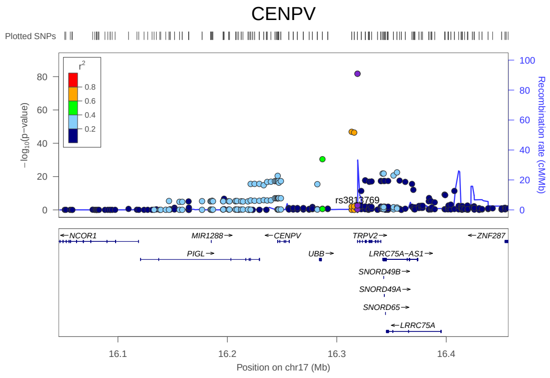


(18) pQTL of CENPV

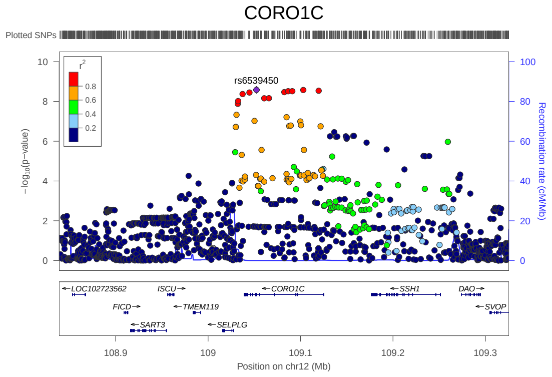


(19) eQTL of CORO1C

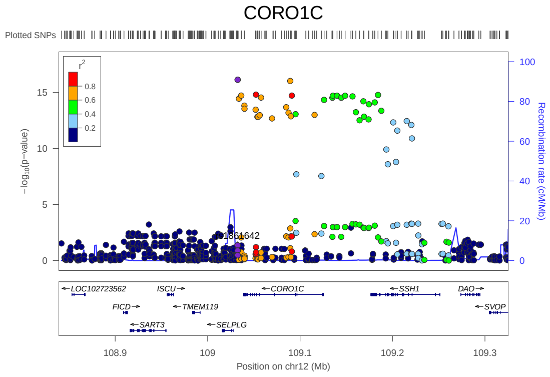


(20) pQTL of CORO1C

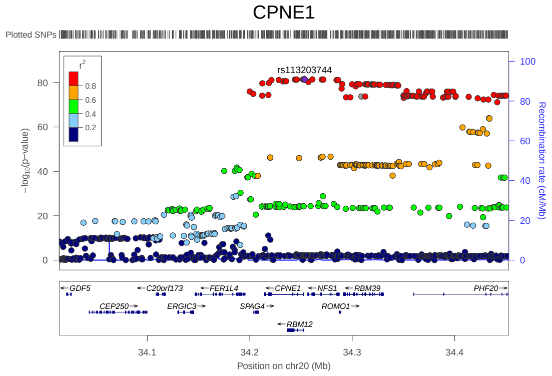


(21) eQTL of CPNE1

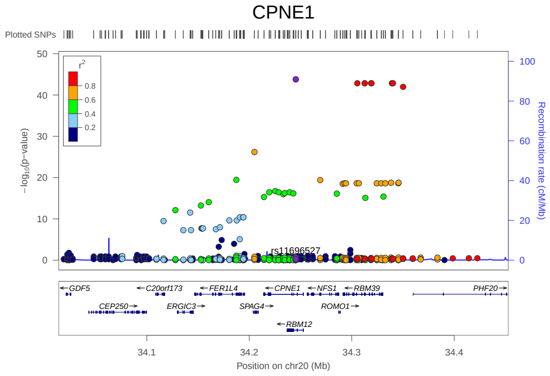


(22) pQTL of CPNE1

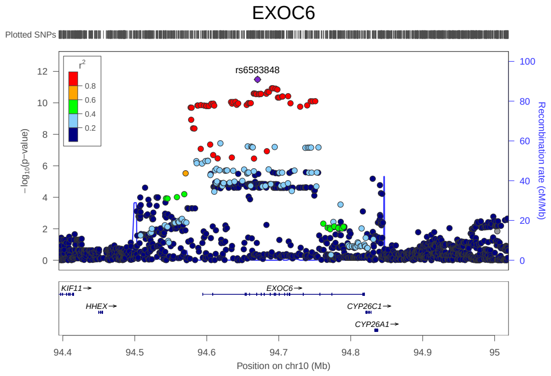


(23) eQTL of EXOC6

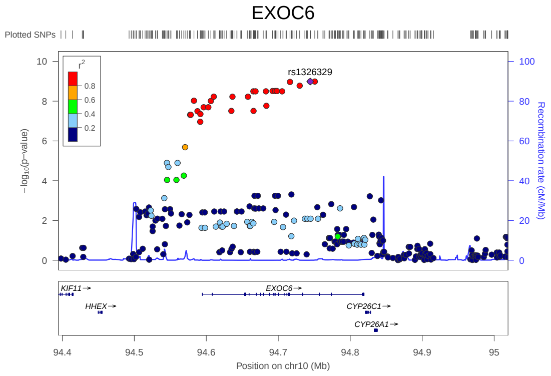


(24) pQTL of EXOC6

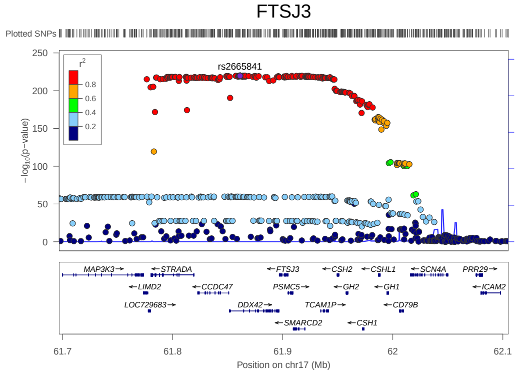


(25) eQTL of FTSJ3

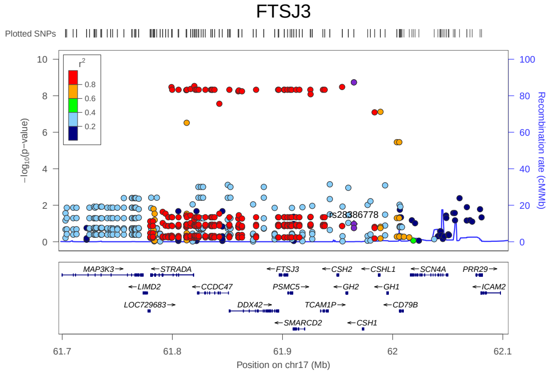


(26) pQTL of FTSJ3

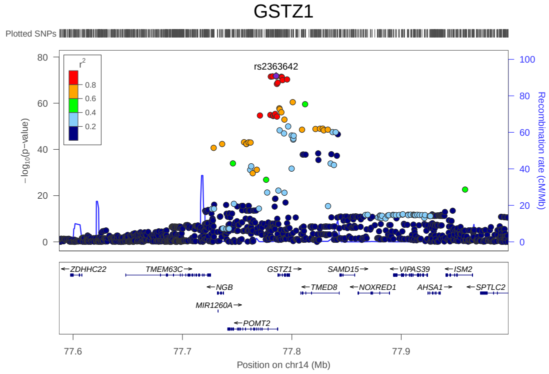


(27) eQTL of GSTZ1

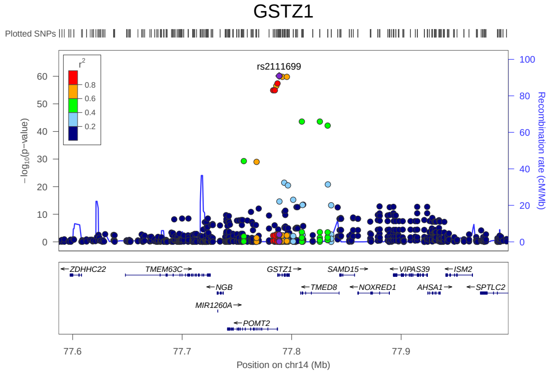


(28) pQTL of GSTZ1

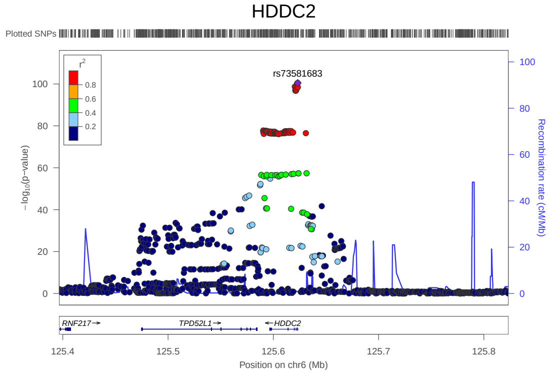


(29) eQTL of HDDC2

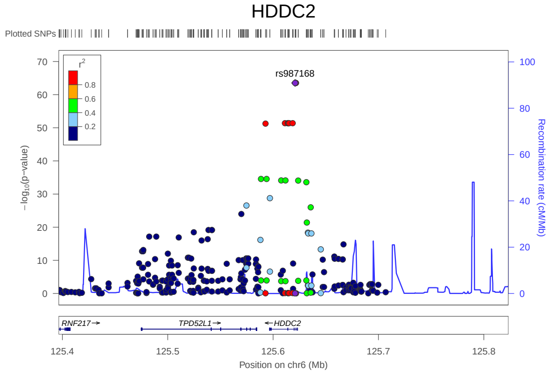


(30) pQTL of HDDC2

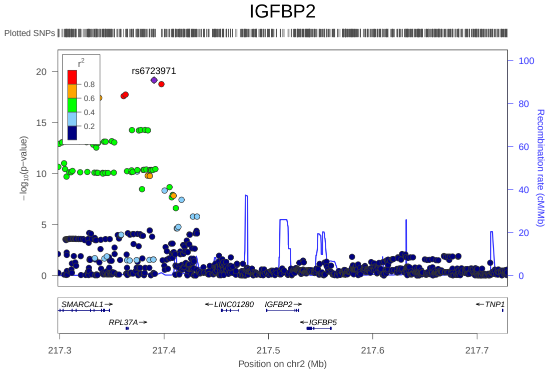


(31) eQTL of IGFBP2

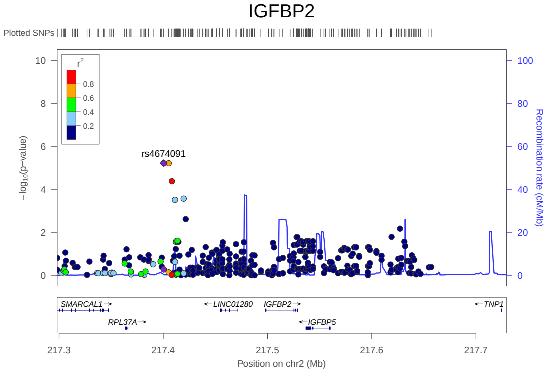


(32) pQTL of IGFBP2

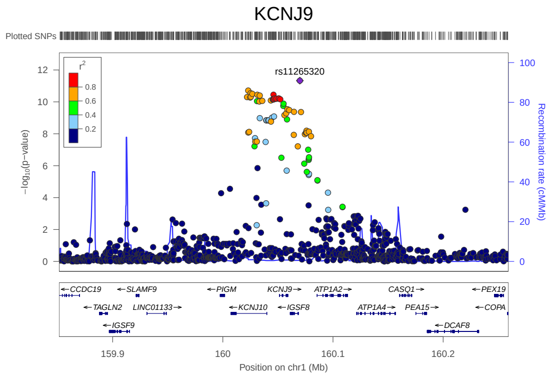


(33) eQTL of KCNJ9

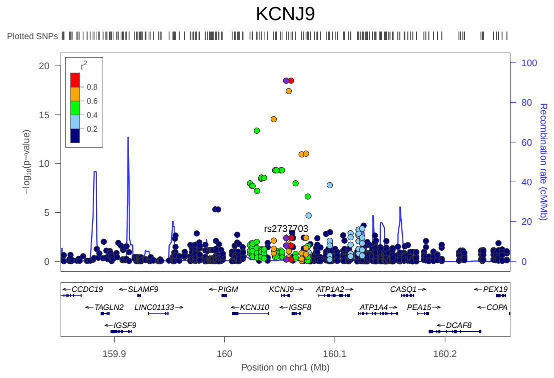


(34) pQTL of KCNJ9

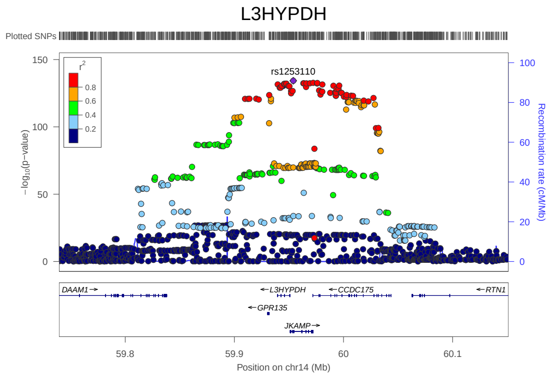


(35) eQTL of L3HYPDH

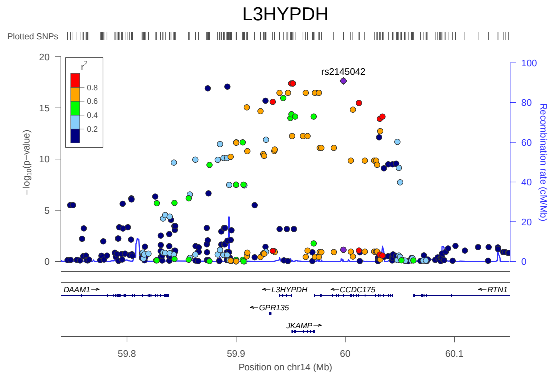


(36) pQTL of L3HYPDH

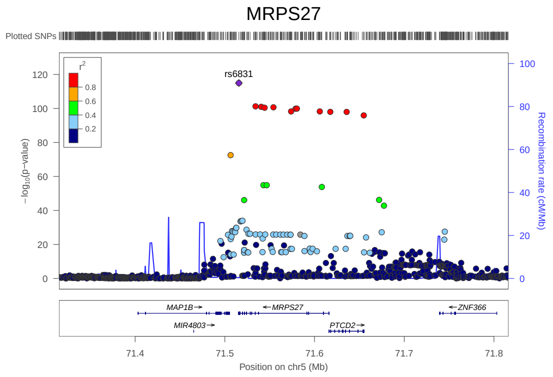


(37) eQTL of MRPS27

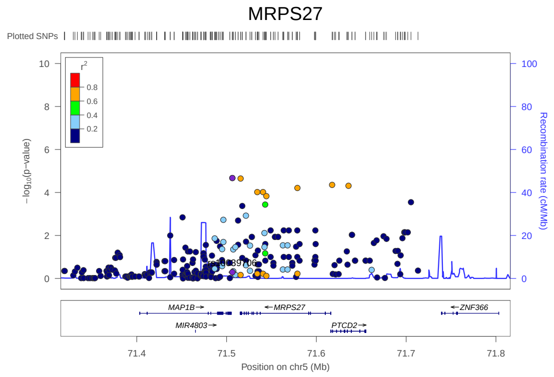


(38) pQTL of MRPS27

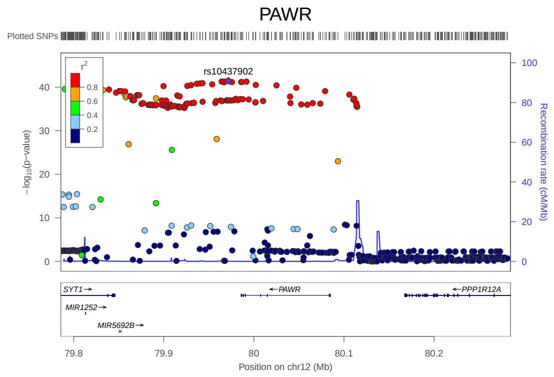


(39) eQTL of PAWR

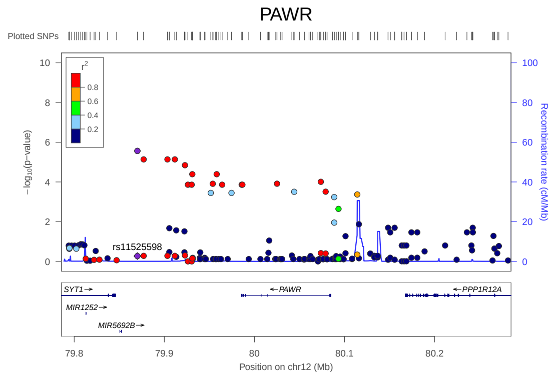


(40) pQTL of PAWR

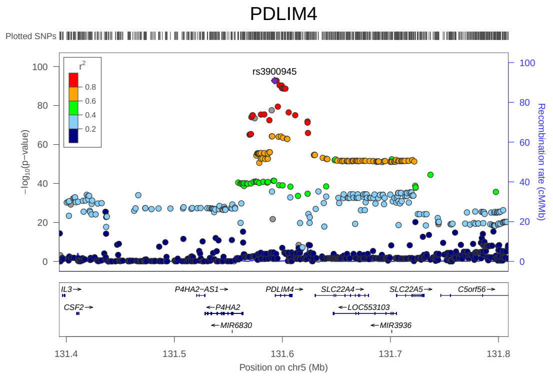


(41) eQTL of PDLIM4

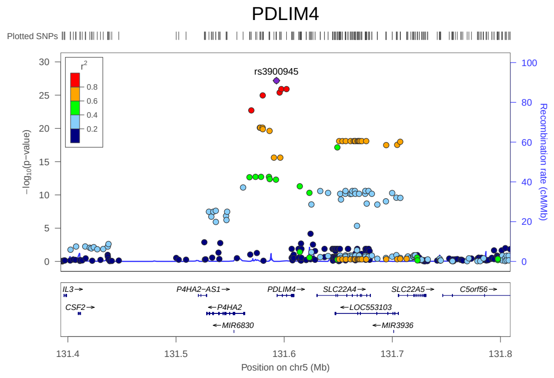


(42) pQTL of PDLIM4

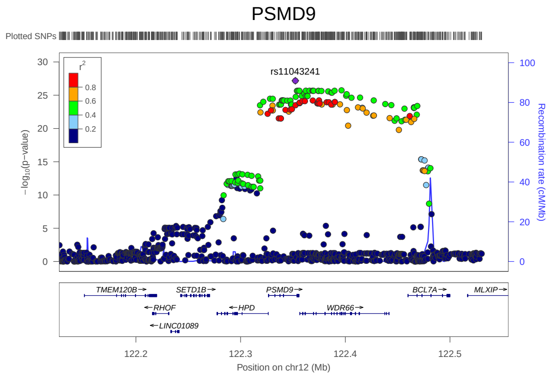


(43) eQTL of PSMD9

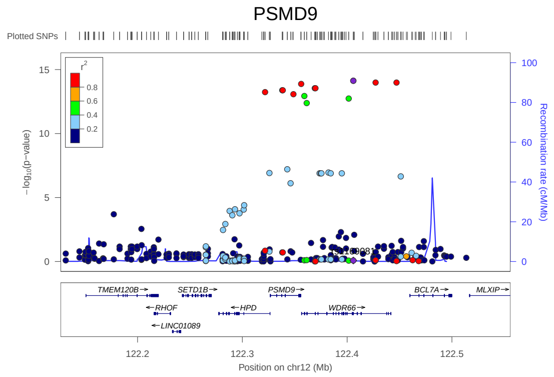


(44) pQTL of PSMD9

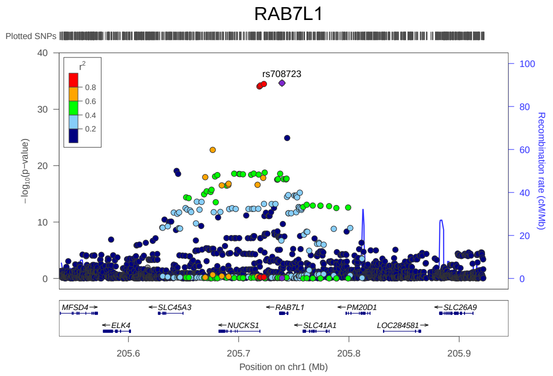


(45) eQTL of RAB7L1

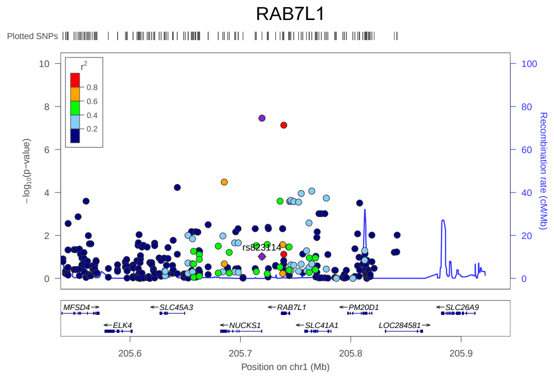


(46) pQTL of RAB7L1

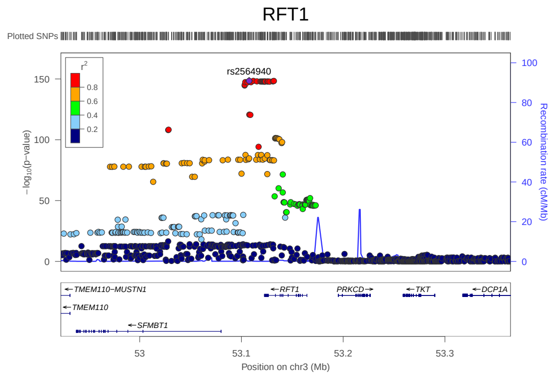


(47) eQTL of RFT1

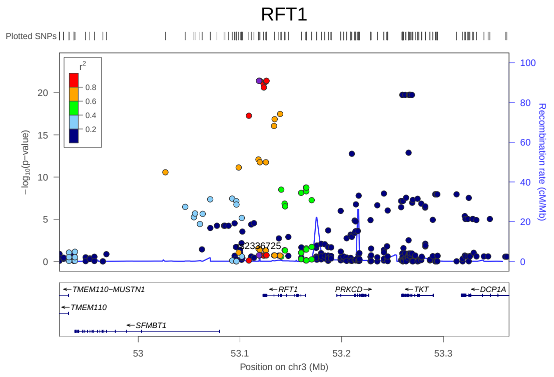


(48) pQTL of RFT1

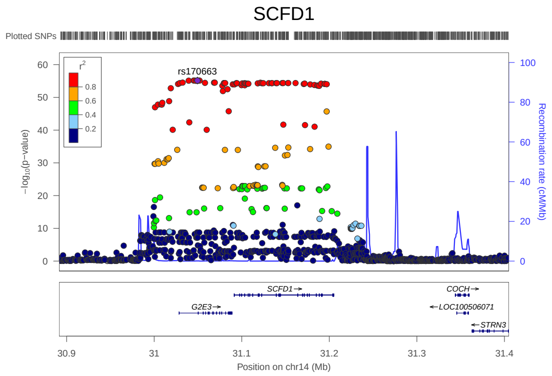


(49) eQTL of SCFD1

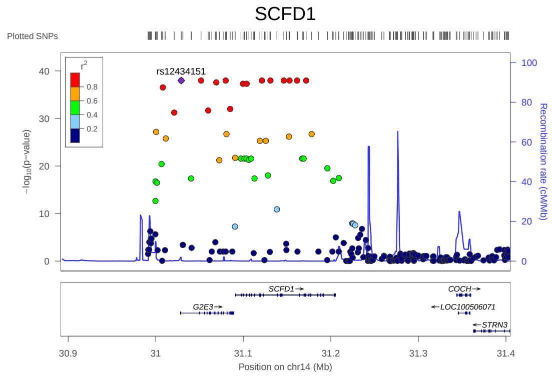


(50) pQTL of SCFD1

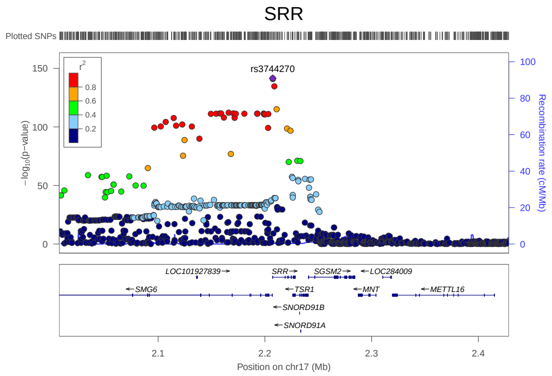


(51) eQTL of SRR

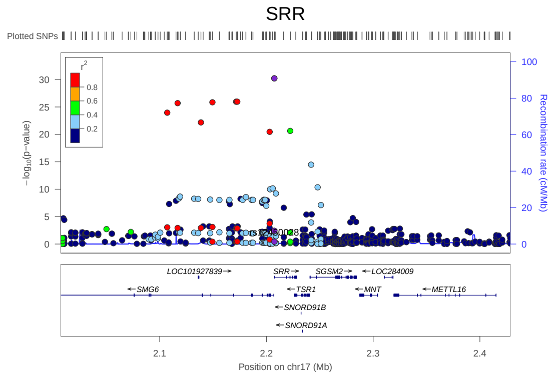


(52) pQTL of SRR

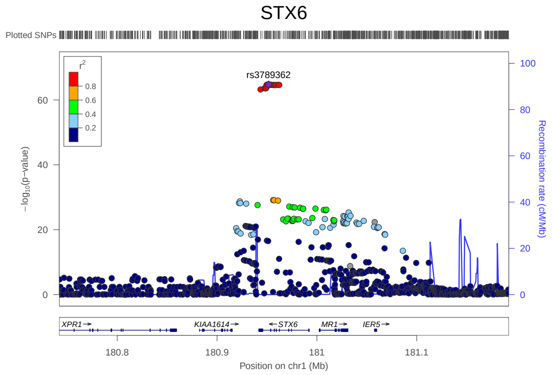


(53) eQTL of STX6

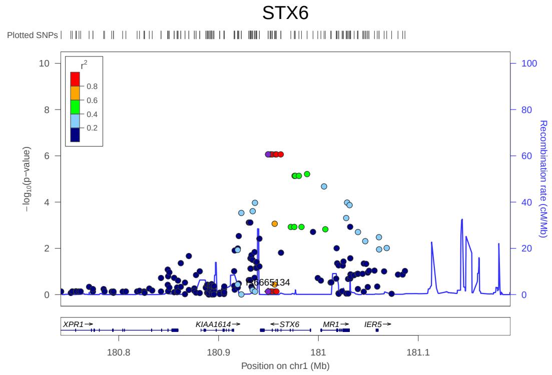


(54) pQTL of STX6

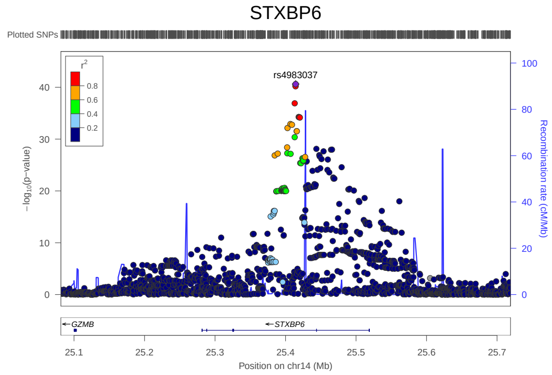


(55) eQTL of STXBP6

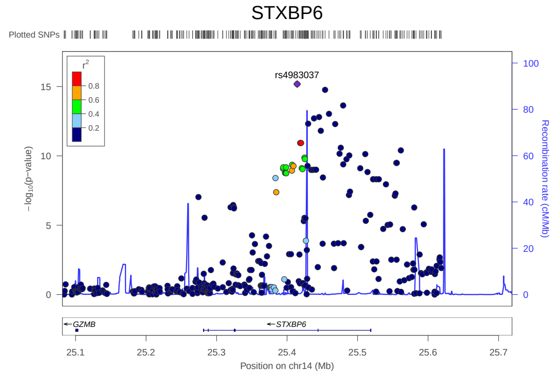


(56) pQTL of STXBP6

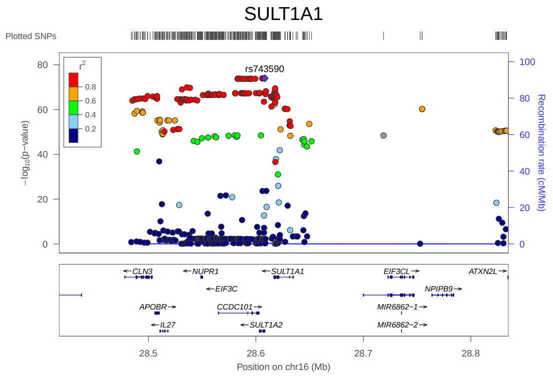


(57) eQTL of SULT1A1

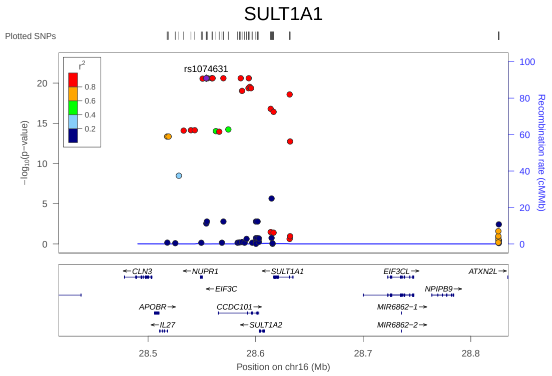


(58) pQTL of SULT1A1

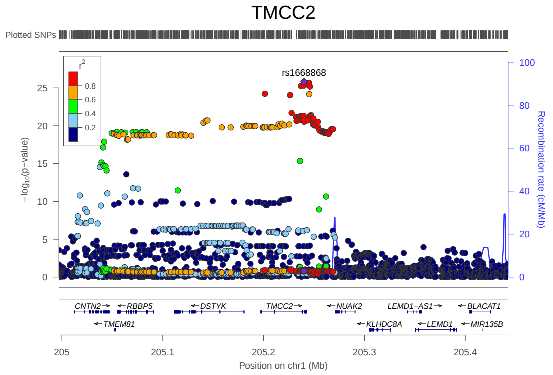


(59) eQTL of TMCC2

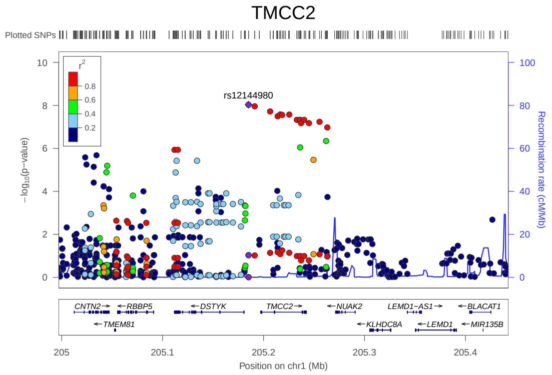


(60) pQTL of TMCC2

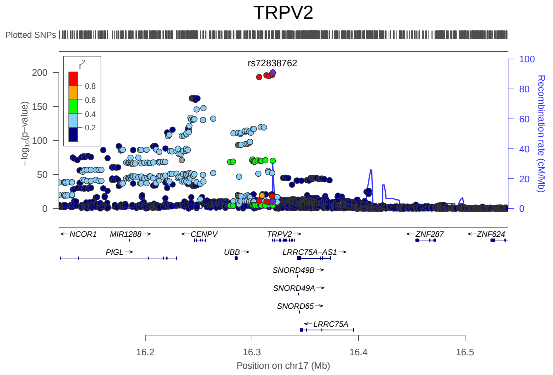


(61) eQTL of TRPV2

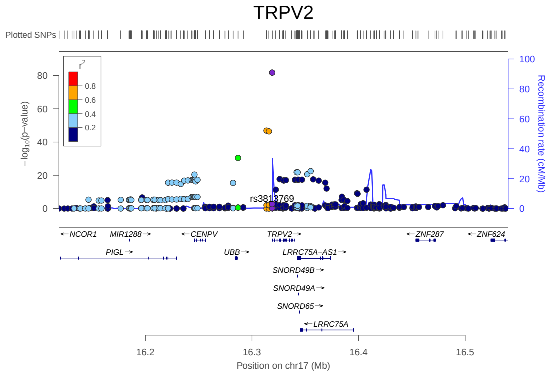


(62) pQTL of TRPV2

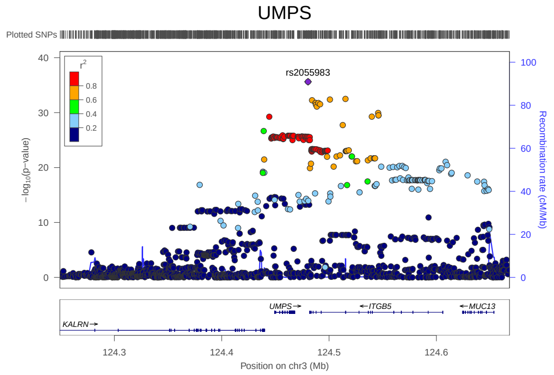


(63) eQTL of UMPS



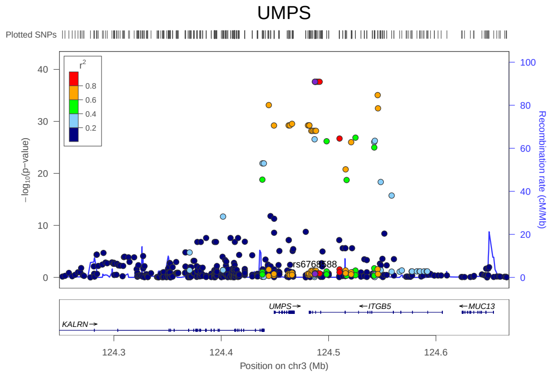


(64) pQTL of UMPS
